# Atrial Cardiomyopathy Refines Cardiovascular Mortality Risk Across Cardiometabolic Risk Factor Burden

**DOI:** 10.64898/2026.09.09.26362666

**Authors:** Asem M. Mohsen, Moustafa Elnewishy, Tarek Zaho, Patrick Cheon, Brian C. Boursiquot, Parag A. Chevli, Richard Kazibwe, Prashant D. Bhave, Elsayed Z. Soliman

## Abstract

**Background:** Electrocardiographic markers of atrial cardiomyopathy (AtCM) are associated with adverse cardiovascular outcomes. Whether AtCM further refines cardiovascular disease (CVD) mortality risk across increasing cardiometabolic risk factor (CMRF) burden is unclear.

**Methods:** We analyzed 7,083 adults free of baseline CVD. AtCM was defined by the presence of ≥1 ECG marker: prolonged P-wave duration ≥120 ms in lead II, abnormal P-wave axis outside 0°–75°, or deep terminal negativity of the P wave in V1 <−100 μV. CMRF burden was defined as the presence of 0, 1, or ≥2 of hypertension, diabetes, and obesity. CVD mortality was ascertained through December 31, 2006.

**Results:** Among 7,083 participants (mean age 58 years and 48% men), CVD mortality rates increased progressively across the six combined exposure groups. In a multivariable adjusted Cox proportional hazard analysis, AtCM identified higher-risk subgroups within each CMRF burden category, with risk increasing progressively across combined exposure groups and highest among participants with ≥2 risk factors and AtCM (HR 2.25, 95% CI 1.75–2.87). In secondary analyses, a similar pattern was observed for individual CMRFs, with the combination of AtCM and hypertension, diabetes, or obesity conferring the highest risk compared with participants with neither condition (HR (95% CI): 1.58 (1.31–1.89); 2.08 (1.62–2.68); and 1.47 (1.19–1.82), respectively).

**Conclusions:** ECG-defined AtCM refined CVD mortality risk across increasing CMRF burden. Individuals with both AtCM and multiple CMRFs had the highest risk. These findings support the potential role of ECG-based AtCM assessment as a scalable approach to improve cardiovascular risk stratification among individuals with CMRFs.

## Introduction

Hypertension, diabetes and obesity are highly prevalent cardiometabolic risk factors that commonly coexist and are leading causes of cardiovascular morbidity and mortality worldwide. [1,2] Beyond their established associations with adverse cardiovascular outcomes, they are increasingly recognized as important drivers of atrial cardiomyopathy (AtCM), a spectrum of structural, electrical, and functional atrial abnormalities that can precede clinically apparent atrial fibrillation (AF) and related cardiovascular complications, including ischemic stroke, heart failure, and mortality. [3–7]

Electrocardiographic (ECG) markers of atrial remodeling; prolonged P-wave duration, abnormal P-wave axis, and deep terminal negativity of the P wave in lead V1, provide a simple, noninvasive means of identifying AtCM. [8,9] Recent studies also suggest that increasing burden of these ECG abnormalities is associated with progressively higher cardiovascular risk in a dose response manner. [10]

Cardiometabolic risk factors and AtCM are closely linked through multiple biological pathways. Hypertension leads to atrial pressure overload and interstitial fibrosis through continuous mechanical stress on myocardium; diabetes drives oxidative stress, autonomic and myocardial dysfunction; and obesity is associated with epicardial adipose tissue expansion, local inflammation, and atrial structural remodeling. [11–13]

Prior studies have largely examined individual cardiometabolic risk factors or ECG markers of AtCM and their associations with cardiovascular risk in isolation. However, it remains unclear whether AtCM further stratifies cardiovascular mortality risk across different levels of cardiometabolic burden. Clarifying this relationship could improve risk stratification among the large population with cardiometabolic disease using simple ECG markers of AtCM.

Therefore, we examined the joint associations of cardiometabolic risk factor (CMRF) burden and AtCM with cardiovascular mortality among participants free of baseline cardiovascular disease in the Third National Health and Nutrition Examination Survey (NHANES III). We hypothesized that AtCM would identify subgroups at disproportionately higher cardiovascular mortality risk within each stratum of CMRF burden.

## Methods

### Study Population

We analyzed data from the Third National Health and Nutrition Examination Survey (NHANES III), a nationally representative survey of the U.S. civilian population conducted by the National Center for Health Statistics (NCHS) between 1988 and 1994, which included standardized interviews, physical examinations, laboratory testing, and resting 12-lead electrocardiograms (ECGs). All participants provided written informed consent, and the study protocol was approved by the NCHS Institutional Review Board. Detailed descriptions of the survey design have been published previously. [14]

The analytic cohort included adults aged ≥40 years with available ECG data who were in sinus rhythm and free of clinically reported CVD and ECG evidence of myocardial infarction.

Participants with missing data on cardiometabolic risk factors, AtCM variables, covariates, or mortality follow-up were excluded, as were participants with body mass index (BMI) <18.5 kg/m², to minimize confounding related to underweight status and frailty. The final analytic cohort comprised 7,083 participants.

## Atrial Cardiomyopathy

AtCM was defined using previously established ECG markers: P-wave duration ≥120 ms in lead II, abnormal P-wave axis outside 0°–75°, and deep terminal negativity of the P wave in lead V1 (DTNPV1) <−100 μV. [15–17] AtCM presence was defined as ≥1 marker.

## Cardiometabolic Risk Factors

Hypertension was defined as systolic blood pressure ≥140 mmHg, diastolic blood pressure ≥90 mmHg, or antihypertensive medication use. Diabetes was defined by self-reported physician diagnosis, glucose-lowering medication or insulin use, or elevated glucose consistent with diabetes. Obesity was defined as BMI ≥30 kg/m². A CMRF burden score summed the presence of these three conditions (0, 1, or ≥2), and combined exposure groups were formed by crossing burden category with AtCM status.

## Outcome Ascertainment

The primary outcome was cardiovascular disease (CVD) mortality, ascertained from National Death Index–linked mortality files and underlying cause-of-death classification, with follow-up from the NHANES III examination date to death or December 31, 2006.

## Covariates

Covariates were selected a priori based on established associations with cardiovascular mortality. Age, sex, race/ethnicity, and smoking status were obtained by standardized interview. Total cholesterol, high-density lipoprotein (HDL) cholesterol, and serum creatinine were measured using standardized laboratory methods.

## Statistical Analysis

Baseline characteristics were compared across the six combined CMRF burden/AtCM groups using one-way analysis of variance for continuous variables and chi-square tests for categorical variables. Continuous variables are presented as mean ± standard deviation, and categorical variables as counts (percentages).

CVD mortality incidence rates were calculated using person-time methods and expressed per 1,000 person-years across the six combined exposure groups. Kaplan–Meier curves were generated to illustrate cumulative CVD mortality across these groups using the product-limit method.

Associations between the combined CMRF burden/AtCM groups and CVD mortality were evaluated using Cox proportional hazards regression, with participants having no CMRF and no AtCM as the reference group. Models were constructed sequentially: Model 1 adjusted for age, sex, race/ethnicity, education, and income; Model 2 additionally adjusted for current smoking, total cholesterol, HDL cholesterol, and serum creatinine. Model 2 was designated the primary model. As a sensitivity analysis, an additional model (Model 3) further adjusted for major and minor ECG abnormalities to assess whether associations reflected broader ECG- detected cardiac disease rather than atrial substrate specifically. Multiplicative interaction between AtCM and CMRF burden was evaluated by including their product term in the primary Cox model.

Secondary analyses examined individual combinations of hypertension, diabetes, and obesity with AtCM status, using the same covariate adjustment as the primary model (Model 2). All statistical tests were two-sided, and a P value <0.05 was considered statistically significant. Analyses were performed using SAS version 9.4 (SAS Institute Inc., Cary, North Carolina).

## Results

Among 7,083 participants free of baseline CVD and in sinus rhythm, mean age was 58 years and 48% were men. Participants were distributed across six combined CMRF burden/AtCM groups. Participants with greater CMRF burden were generally older, had a higher BMI, higher systolic and diastolic blood pressure and generally poorer cardiovascular risk profile. Within each CMRF category, participants with AtCM were older and had a higher prevalence of current smoking and ECG abnormalities than those without AtCM **(Table 1).**

**Table 1.** Baseline Characteristics by Cardiometabolic Risk Factor Burden and AtCM Status

| Characteristic* | No CMRFs |  | 1 CMRF |  | ≥ 2 CMRFs |  | p-value |
| --- | --- | --- | --- | --- | --- | --- | --- |
|  | – AtCM | + AtCM | – AtCM | + AtCM | – AtCM | + AtCM |  |
| N | 1877 | 1470 | 1370 | 1087 | 729 | 550 |  |
| Age, years | 55.9 ± 13.1 | 60.0 ± 13.9 | 57.6 ± 13.1 | 63.1 ± 13.4 | 58.7 ± 11.7 | 61.3 ± 11.8 | <0.001 |
| Male sex, n (%) | 927 (49.4%) | 811 (55.2%) | 554 (40.4%) | 551 (50.7%) | 245 (33.6%) | 225 (40.9%) | <0.001 |
| Education, years | 10.8 ± 4.5 | 10.8 ± 4.4 | 10.3 ± 4.3 | 10.2 ± 4.3 | 9.8 ± 4.2 | 10.0 ± 4.2 | <0.001 |
| Follow-up, years, median (IQR) | 14.5 (12.7–16.2) | 13.9 (11.1–16.0) | 13.9 (12.4–15.9) | 13.3 (9.3–15.5) | 13.5 (11.7–15.5) | 13.2 (9.2–15.3) | <0.001 |
| Race/ethnicity |  |  |  |  |  |  |  |
| Non-Hispanic White | 972 (51.8%) | 832 (56.6%) | 621 (45.3%) | 516 (47.5%) | 280 (38.4%) | 231 (42.0%) | <0.001 |
| Non-Hispanic Black | 314 (16.7%) | 319 (21.7%) | 312 (22.8%) | 324 (29.8%) | 199 (27.3%) | 197 (35.8%) | <0.001 |
| Mexican American | 497 (26.5%) | 257 (17.5%) | 377 (27.5%) | 215 (19.8%) | 216 (29.6%) | 115 (20.9%) | <0.001 |
| Other race/ethnicity | 94 (5.0%) | 62 (4.2%) | 60 (4.4%) | 32 (2.9%) | 34 (4.7%) | 7 (1.3%) | 0.001 |
| Income <\$20,000, n (%) | 713 (38.0%) | 646 (43.9%) | 658 (48.0%) | 561 (51.6%) | 406 (55.7%) | 274 (49.8%) | <0.001 |
| Current smoker, n (%) | 432 (23.0%) | 449 (30.5%) | 242 (17.7%) | 261 (24.0%) | 107 (14.7%) | 93 (16.9%) | <0.001 |
| Systolic BP, mmHg | 124.8 ± 16.9 | 126.9 ± 17.9 | 134.3 ± 18.5 | 139.1 ± 20.9 | 140.6 ± 19.5 | 142.0 ± 19.3 | <0.001 |
| Diastolic BP, mmHg | 74.5 ± 9.0 | 74.0 ± 9.5 | 78.3 ± 10.2 | 77.7 ± 11.3 | 79.3 ± 11.1 | 79.1 ± 11.3 | <0.001 |
| BMI, kg/m² | 25.3 ± 2.7 | 24.3 ± 3.0 | 29.4 ± 5.1 | 28.2 ± 5.4 | 33.3 ± 4.8 | 34.1 ± 6.1 | <0.001 |
| Total cholesterol, mg/dL | 208.6 ± 56.1 | 204.4 ± 58.5 | 212.6 ± 55.8 | 212.8 ± 60.0 | 214.1 ± 64.7 | 211.2 ± 62.0 | <0.001 |
| HDL cholesterol, mg/dL | 49.8 ± 18.2 | 52.4 ± 20.7 | 47.4 ± 17.0 | 48.8 ± 19.1 | 45.3 ± 19.6 | 45.4 ± 17.9 | <0.001 |
| Triglycerides, mg/dL | 143.2 ± 106.1 | 126.2 ± 87.9 | 166.9 ± 132.7 | 154.3 ± 115.2 | 202.8 ± 212.4 | 179.8 ± 146.4 | <0.001 |
| Hemoglobin A1c, % | 5.3 ± 1.2 | 5.3 ± 1.1 | 5.7 ± 1.6 | 5.7 ± 1.5 | 6.4 ± 2.2 | 6.3 ± 2.0 | <0.001 |
| Serum creatinine, mg/dL | 1.0 ± 0.3 | 1.0 ± 0.3 | 1.0 ± 0.4 | 1.1 ± 0.4 | 1.1 ± 0.8 | 1.1 ± 0.9 | <0.001 |
| Antihypertensive medication use, n (%) | 0 (0.0%) | 0 (0.0%) | 363 (26.5%) | 423 (38.9%) | 422 (57.9%) | 342 (62.2%) | <0.001 |
| Hypertension, n (%) | 0 (0.0%) | 0 (0.0%) | 626 (45.7%) | 639 (58.8%) | 644 (88.3%) | 505 (91.8%) | <0.001 |
| Diabetes, n (%) | 0 (0.0%) | 0 (0.0%) | 142 (10.4%) | 100 (9.2%) | 301 (41.3%) | 209 (38.0%) | <0.001 |
| Obesity, n (%) | 0 (0.0%) | 0 (0.0%) | 602 (43.9%) | 348 (32.0%) | 617 (84.6%) | 471 (85.6%) | <0.001 |
| Lipid-lowering medication use, n (%) | 32 (1.7%) | 23 (1.6%) | 53 (3.9%) | 43 (4.0%) | 36 (4.9%) | 26 (4.7%) | <0.001 |
| Major ECG abnormality, n (%) | 188 (10.0%) | 180 (12.2%) | 166 (12.1%) | 187 (17.2%) | 109 (15.0%) | 117 (21.3%) | <0.001 |
| Minor ECG abnormality, n (%) | 315 (16.8%) | 285 (19.4%) | 283 (20.7%) | 305 (28.1%) | 196 (26.9%) | 168 (30.5%) | <0.001 |
| Prolonged P-wave duration, n (%) | 0 (0.0%) | 674 (45.9%) | 0 (0.0%) | 713 (65.6%) | 0 (0.0%) | 423 (76.9%) | <0.001 |
| Abnormal P-wave axis, n (%) | 0 (0.0%) | 964 (65.6%) | 0 (0.0%) | 489 (45.0%) | 0 (0.0%) | 160 (29.1%) | <0.001 |
| Abnormal DTNPV1, n (%) | 0 (0.0%) | 63 (4.3%) | 0 (0.0%) | 57 (5.2%) | 0 (0.0%) | 38 (6.9%) | <0.001 |
| CVD death, n (%) | 161 (8.6%) | 188 (12.8%) | 151 (11.0%) | 194 (17.8%) | 112 (15.4%) | 114 (20.7%) | <0.001 |

AtCM, atrial cardiomyopathy; –, absent; +, present; CMRF, cardiometabolic risk factor; BP, blood pressure; BMI, body mass index; HDL-cholesterol, high-density lipoprotein cholesterol; DTNPV1, deep terminal negativity in V1; CVD, cardiovascular disease \*Continuous variables are mean ± SD except follow-up, shown as median (IQR); categorical variables are n (%).

During a median follow-up of 13.9 years (IQR, 12.2–15.8 years), 920 CVD deaths occurred. CVD mortality rates increased progressively across combined exposure groups, from 6.27 per 1,000 person-years among participants with no CMRF and no AtCM to 17.27 per 1,000 person-years among those with ≥2 CMRF and AtCM present. In a multivariable adjusted Cox proportional hazard analysis, this pattern persisted, with the risk increasing progressively with increasing CMRF burden and highest among participants with ≥2 risk factors and AtCM (HR 2.25, 95% CI 1.75–2.87). Notably, AtCM was consistently associated with additional risk beyond CMRF burden alone within each stratum and did not show statistical significance when present alone in absence of risk factor **(Figure 1)**. There was no significant multiplicative interaction between AtCM and CMRF burden (P = 0.883) In the sensitivity model (Model 3) additionally adjusted for major and minor ECG abnormalities, these associations were nearly unchanged **(Table 2).** Kaplan–Meier curves further demonstrated progressively lower survival probabilities across increasing cardiometabolic risk factor burden categories and AtCM presence **(Figure 2)**.

**Figure 1.**
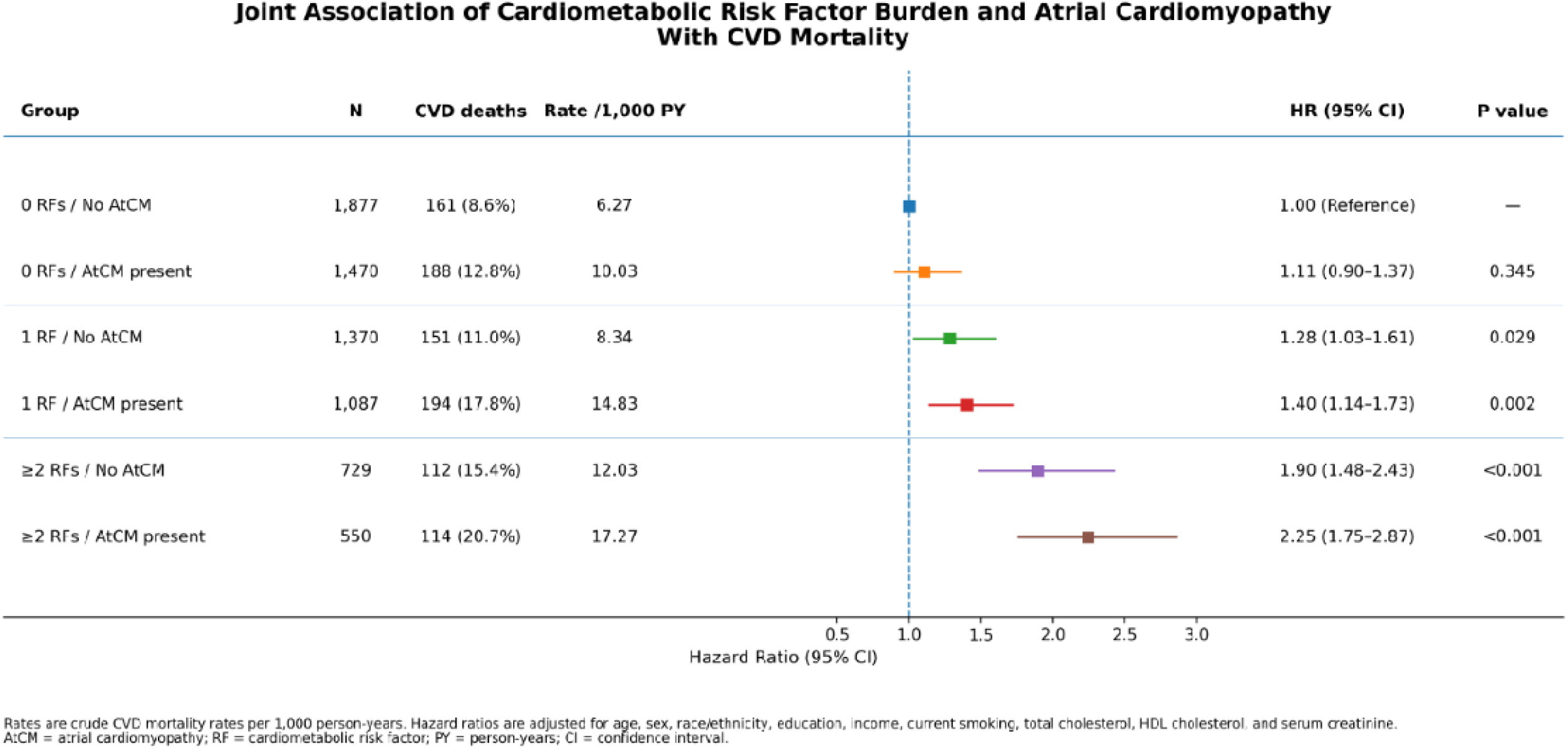
CVD Mortality Rates and Adjusted Hazard Ratios According to Cardiometabolic Risk Factor Burden and AtCM Crude CVD mortality rates and fully adjusted hazard ratios are shown for the six combined CMRF burden/AtCM groups. Hazard ratios are adjusted for age, sex, race/ethnicity, education, income, current smoking, total cholesterol, HDL cholesterol, and serum creatinine. AtCM absent/no risk factor is the reference group for all groups. CMRF, cardiometabolic risk factor; AtCM, atrial cardiomyopathy; CVD, cardiovascular disease; CI, confidence interval.

**Figure 2.**
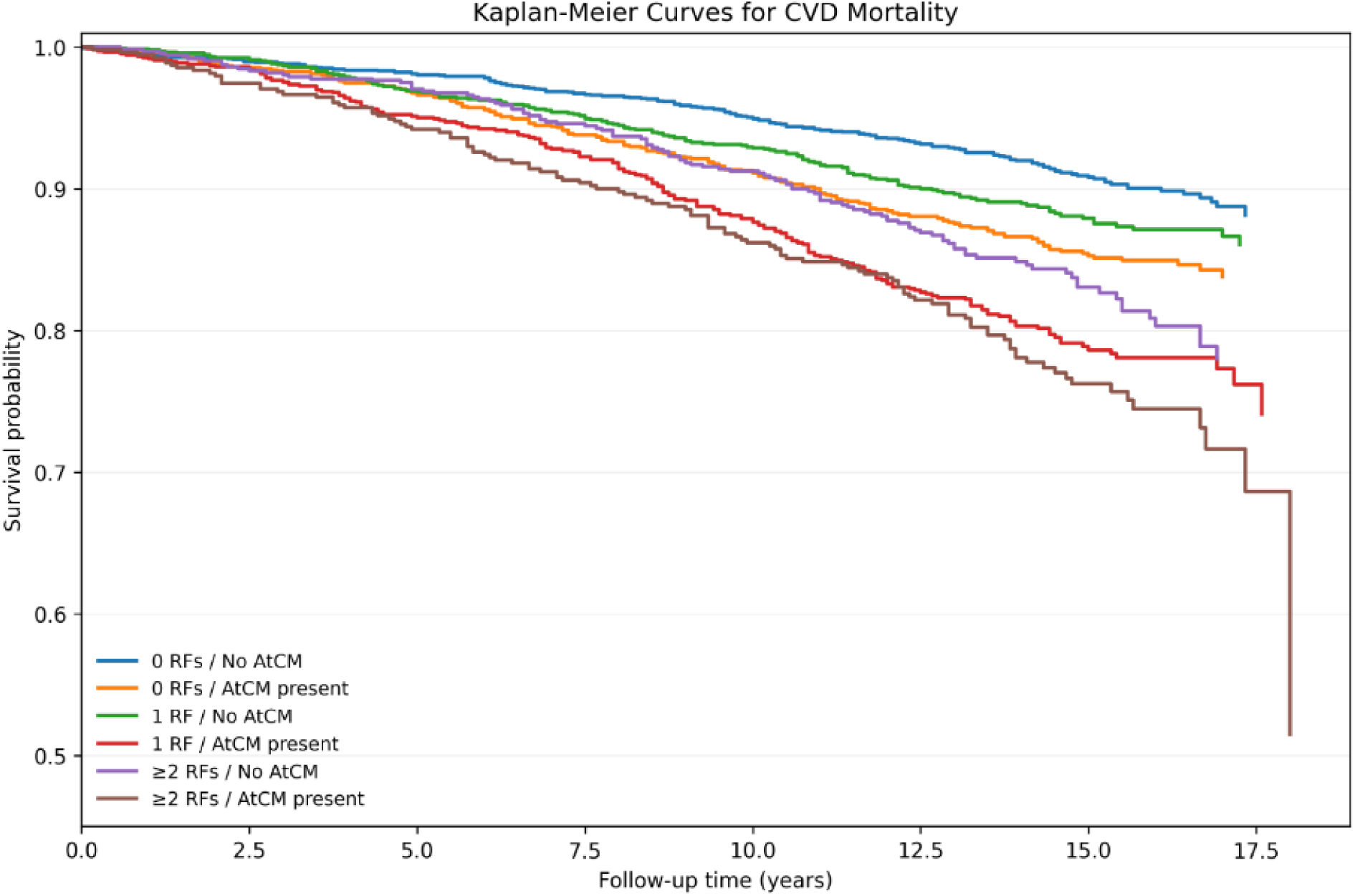
Kaplan-Meier Curves for CVD-Free Survival by AtCM Status and Cardiometabolic Risk Factor Burden Kaplan–Meier survival curves for CVD death are shown for the six combined groups defined by CMRF burden (0, 1, or ≥2 of hypertension, diabetes, and obesity) and AtCM status (presence of ≥1 marker vs. absence), with CVD death as the event. CMRF, cardiometabolic risk factor; AtCM, atrial cardiomyopathy; CVD, cardiovascular disease.

**Table 2.** CVD Mortality Rates and HRs According to Cardiometabolic Risk Factor Burden and AtCM Status

| AtCM Status/CMRF Burden | HR (95% CI) |  |  | Mortality<br>Rate |
| --- | --- | --- | --- | --- |
|  | Model 1□ | Model 2† | Model 3* |  |
| AtCM absent/no risk factor | 1.00 (Reference) | 1.00 (Reference) | 1.00 (Reference) | 6.27 |
| AtCM present/ no risk factor | 1.17 (0.95–1.44) | 1.11 (0.90–1.37) | 1.11 (0.90–1.37) | 10.03 |
| AtCM absent/ 1 risk factor | 1.27 (1.02–1.59) | 1.28 (1.03–1.61) | 1.26 (1.01–1.58) | 8.34 |
| AtCM present / 1 risk factor | 1.43 (1.16–1.77) | 1.40 (1.14–1.73) | 1.36 (1.10–1.68) | 14.83 |
| AtCM absent/ ≥2 risk factors | 1.86 (1.46–2.38) | 1.90 (1.48–2.43) | 1.86 (1.45–2.39) | 12.03 |
| AtCM present/ ≥2 risk factors | 2.23 (1.74–2.84) | 2.25 (1.75–2.87) | 2.13 (1.66–2.73) | 17.27 |

Reference group: 0 cardiometabolic risk factors and no atrial cardiomyopathy. □Model 1: adjusted for age, sex, race/ethnicity, education, and income. †Model 2: Model 1 plus smoking, total cholesterol, HDL cholesterol, and serum creatinine (Primary model). \*Model 3: Model 2 plus major and minor ECG abnormalities (sensitivity model). Mortality rate per 1000 person years RF, Risk factor; AtCM, atrial cardiomyopathy; CI, confidence interval; HR, hazard ratio

In secondary analyses evaluating each cardiometabolic risk factor individually and in combination with AtCM, similar patterns were observed. Compared with participants with neither condition, CVD mortality risk was highest among those with both AtCM and hypertension (HR 1.58, 95% CI 1.31–1.89), diabetes (HR 2.08, 95% CI 1.62–2.68), or obesity (HR 1.47, 95% CI 1.19–1.82). Corresponding HRs for the risk factor without AtCM were 1.42 (95% CI 1.17–1.72), 1.71 (95% CI 1.33–2.19), and 1.29 (95% CI 1.04–1.60), respectively, whereas AtCM in the absence of the respective risk factor was not significantly associated with CVD mortality. **(Table 3)**

**Table 3.**
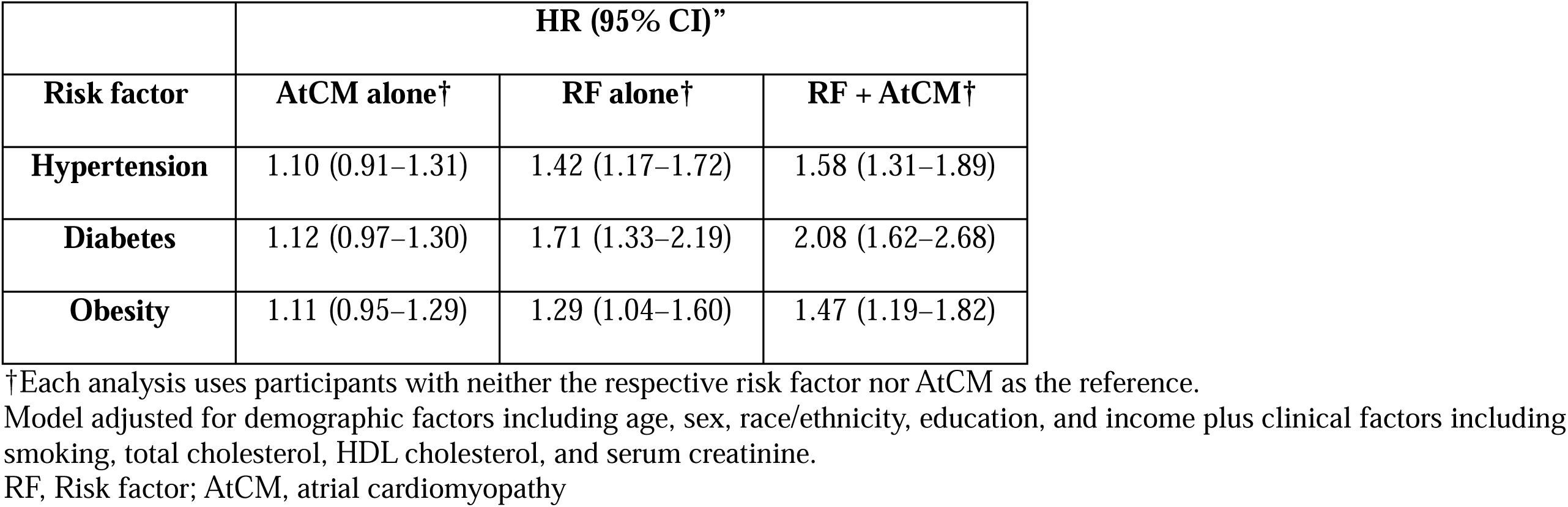
Cardiovascular Mortality Risk According to Individual Cardiometabolic Risk Factors and AtCM.

## Discussion

In this community-based cohort of adults free of baseline CVD, increasing CMRF burden was associated with progressively higher CVD mortality risk. More importantly, AtCM presence was consistently associated with additional higher risk beyond CMRF burden alone within each stratum, identifying higher-risk subgroups within each CMRF burden category. Participants with both ≥2 cardiometabolic risk factors and AtCM had the highest mortality rates, conferring more than a twofold higher adjusted risk of CVD death compared with participants with neither. When analyzed individually, a similar pattern was observed across all three risk factors, with the combination of AtCM and hypertension, diabetes, or obesity conferring greater risk than either the risk factor or AtCM alone, and AtCM in the absence of the respective risk factor not reaching statistical significance.

Our findings extend prior evidence linking ECG markers of AtCM to adverse CVD outcomes. [6,7,18,19] However, most of these studies have focused on isolated ECG abnormalities or individual risk factors. The present study builds on previous work by demonstrating that AtCM may serve as an important marker of heightened cardiovascular susceptibility across increasing cardiometabolic risk burden.

Hypertension, diabetes and obesity are well-known cardiometabolic risk factors that contribute to atrial structural and electrical remodeling through several interconnected mechanisms. [12,20–22] Hypertension promotes atrial stretch and fibrosis through chronic pressure overload, [20,23] whereas diabetes contributes to metabolic and microvascular dysfunction that may accelerate atrial remodeling. [7,13,24] Obesity is associated with systemic inflammation, epicardial adiposity, and atrial enlargement, all of which may facilitate development of AtCM. [12,25] These mechanisms likely interact synergistically, contributing to progressive atrial remodeling and increased susceptibility to cardiovascular events. [26]

An important finding of this study is that AtCM was associated with higher CVD mortality only among participants with cardiometabolic risk factors. Among participants with one CMRF, the presence of AtCM was associated with higher CVD mortality compared with those without AtCM, and a similar association was observed among participants with ≥2 cardiometabolic risk factors. In contrast, among participants with no cardiometabolic risk factors, the association between AtCM and CVD mortality did not reach statistical significance. These findings suggest that AtCM may be particularly informative among individuals with established cardiometabolic risk factors.

Our findings may have potential clinical implications. ECG-derived P-wave markers of AtCM are inexpensive, widely available, and routinely obtained in clinical practice. Incorporation of AtCM assessment into cardiovascular risk stratification strategies may have potential clinical utility specifically for individuals at higher risks with cardiometabolic risk factors.

Several limitations merit consideration. The observational design precludes inference of causality, and residual confounding cannot be excluded despite multivariable adjustment. Cardiometabolic risk factors and ECG measurements were assessed at a single baseline examination and may have changed over time. ECG-defined AtCM may not fully capture structural and functional atrial abnormalities that can be detected by imaging. Subclinical AF may have been undetected in some participants. Finally, although NHANES III provides a nationally representative sample, the findings may not fully generalize to contemporary populations given temporal changes in cardiometabolic disease management and cardiovascular prevention strategies.

Strengths of this study include the large, community-based racially diverse cohort with long-term mortality follow-up, standardized ECG acquisition and centralized processing enabled robust assessment of AtCM markers, the study included participants free of baseline CVD and restricted to sinus rhythm, reducing confounding by prevalent CVD and clinically recognized arrhythmias. Finally, we evaluated both cumulative cardiometabolic risk burden and individual cardiometabolic risk factors, allowing a comprehensive assessment of the interplay between systemic cardiometabolic dysfunction and AtCM.

## Conclusion

Among US adults free of baseline cardiovascular disease, increasing cardiometabolic risk factor burden was associated with progressively higher cardiovascular mortality risk, and ECG-defined AtCM further identified subgroups at particularly elevated risk within each CMRF burden category. The coexistence of AtCM with hypertension, diabetes, obesity, or greater cumulative cardiometabolic risk burden was consistently associated with adverse cardiovascular outcomes. These findings support the potential role of ECG-based AtCM assessment as a scalable approach to improve cardiovascular risk stratification among individuals with cardiometabolic risk factors.

## Data Availability

All data produced are available online at https://wwwn.cdc.gov/nchs/nhanes/nhanes3/

## Statements and Declarations

### Data Availability

The data supporting this study are openly available in NHANES III at https://wwwn.cdc.gov/nchs/nhanes/default.aspx.

### Ethics Approval

NHANES III was approved by the National Center for Health Statistics (NCHS) Research Ethics Review Board.

### Consent to Participate

Written informed consent was obtained from all participants as part of NHANES III.

## Author Contributions (CRediT)

A.M.M: Writing – original draft, conceptualization and revisions. M.E: Writing – review and editing and data curation. T.Z: Writing – review and editing. P.C: Writing – review and editing. B.C.B: Writing – review and editing. P.A.C: Writing – review and editing. R.K: Writing – review and editing. P.D.B: Writing – review and editing. E.Z.S: Conceptualization, formal analysis, supervision, writing – review and editing.

